# Efficacy and safety of dihydroartemisinin-piperaquine for the treatment of uncomplicated *Plasmodium falciparum* and *Plasmodium vivax* malaria in Northern Papua and Jambi, Indonesia

**DOI:** 10.1101/2020.09.04.20188706

**Authors:** Puji BS Asih, Ismail E Rozi, Farahana K Dewayanti, Suradi Wangsamuda, Syarifah Zulfah, Marthen Robaha, Jonny Hutahaean, Nancy D Anggraeni, Marti Kusumaningsih, Pranti S Mulyani, Elvieda Sariwati, Herdiana H Basri, Maria Dorina G Bustos, Din Syafruddin

## Abstract

Dihydroartemisinin-piperaquine (DHA-PPQ) has been adopted as first-line therapy for uncomplicated falciparum malaria in Indonesia since 2010. The efficacy of DHA-PPQ was evaluated in 2 sentinel sites in Keerom District, Papua and Merangin District, Jambi Provinces from April 2017 to April 2018. Clinical and parasitological parameters were monitored over a 42-day period following the WHO standard *in vivo* protocol and subjects meeting the inclusion criteria were treated with DHA-PPQ once daily for 3 days, administered orally. In Keerom District, 6339 subjects were screened through active and passive cases detection. A total of 114 subjects infected by *P. falciparum* and 83 subjects infected by *P. vivax* agreed to take a part through written informed consent. Kaplan-Meier analysis of microscopy readings and PCR-corrected falciparum cases revealed a 93.1% (95%CI:86.4-97.2) and 97.9% (95%CI:92.7-99.7) DHA-PPQ efficacy, respectively and were classified as Adequate Clinical Parasitological Responses (ACPRs). For vivax malaria, the DHA-PPQ efficacy were 89% (95%CI: 80.2 - 94.9) and 100% (95%CI: 95.1-100) respectively. In Merangin District, 751 subjects were screened and 41 subjects infected by *P. vivax* were recruited. Microscopy reading and PCR-corrected analysis revealed a 97.4% (95%CI:86.2-99.9) and 100% (95%CI: 90.5-100) DHA-PPQ efficacy, respectively. No severe adverse events were found in both sites. In both sites, there was no delay in parasite clearance and no mutations in the PfK13 and PvK12 genes. Of the 6 recurrent *P. falciparum* found, 2 indicated recrudescent and 4 cases were re-infection. Analysis of the PfPM2 gene at day 0 and day of recurrence in recrudescent cases revealed the same single copy number, whereas 3 of the 4 re-infection cases carried 2-3 copy numbers. In conclusion, treatment of falciparum and vivax malaria cases with DHA-PPQ showed a high efficacy and safety. DHA-PPQ regimen is also efficacious against *P. vivax* cases in the absence of primaquine.

## Introduction

In Indonesia, reports to date revealed that Artemisinin Combination Therapies (ACTs), particularly dihydroartemisinin-piperaquine (DHA-PPQ) are still highly effective to treat any human malaria cases. Although certain studies reported few cases of delayed parasite clearance [1], this evidence was not linked to the artemisinin resistance. Subsequent analysis on the cases revealed that the delay may be related to the higher parasite load as the parasite is eventually eliminated by day 7. Therefore, routine monitoring of the therapeutic efficacy of ACTs is essential for making timely changes of treatment policy. It can also help to detect early changes in the parasite susceptibility to antimalarial drugs [2].

Malaria control program in Indonesia has successfully brought down the malaria cases within the last few decades and in 2017, more than half of the district and municipality have been certified as malaria free areas. However, malaria cases remain high in eastern provinces, such as Papua, West Papua, Molucca and East Nusa Tenggara. In Western part of the country, malaria is either eliminated or significantly reduced and only several malaria foci left in Sumatra, Java, Bali and Kalimantan. In 2017 Indonesia reported 261,000 malaria cases with 74% of infections reported from Papua Province [3]. Malaria problem in Indonesia represents a unique archipelago setting that is entirely different with that of Africa. Malaria control program relies on three pillars such as early diagnosis and prompt treatment, provision of LLIN and indoor residual spraying (IRS) [4]. Unfortunately, the health care facilities in remote setting where malaria is highly endemic does not always meet the requirement to provide necessary service to the people. The absence of microscopists and vector control officers reduce the effectiveness of the pillar and also provision of diagnosis and prompt treatment. To avoid the unnecessary antimalarial drug deployment, the Ministry of Health has set a treatment guideline in which antimalarial drug will only be given to laboratory confirmed cases, either by microscopy or rapid diagnostic test (RDT). In Indonesia, follow up of the malaria treatment is rarely done and therefore supervisory treatment has been recommended to ensure that the patients indeed consumed the antimalarial as prescribed. Since 2010, Indonesia recommended DHA-PPQ as first line drug for uncomplicated malaria [5]. This includes all species of human malaria. The side-effects of DHA-PPQ are abdominal pain, asthenia, cough, diarrhea, dizziness, fever, headache, joint and muscle pain, loss of appetite, rush, nausea, vomiting. abdominal discomfort, nausea, headache and dizziness. The objective of this study is to assess the therapeutic efficacy and safety of DHA-PPQ for the treatment of uncomplicated *P. falciparum* and *P. vivax* malaria in Indonesia.

## Methods

### Ethics statement

This study was approved by the Ethics Committee of Research in Health, Medical Faculty of Hasanuddin University, Makassar, Indonesia (No. 663/H4.8.4.5.31/PP36-KOMETIK/2016 and No. 356/H4.8.4.5.31/PP36-KOMETIK/2017). The trial was registered with the clinical trial number ACTRN12616001533482.

### Study Site

The study was conducted in Keerom District and Merangin District from April 2017 – April 2018. The location of the Keerom District in Papua and Merangin District in Jambi Provinces within Indonesia archipelago is shown in Figure 1. The climate is typically tropical in both sites. In Merangin District, Jambi Province the rainy season occurs from December to April, whereas in Keerom District rainy and dry season could not be separated clearly as the rainfall occurred throughout the year. The average temperatures are 18-20°C during the cooler rainy months from December to April and 25-33°C during the dry season.

**Figure 1.**
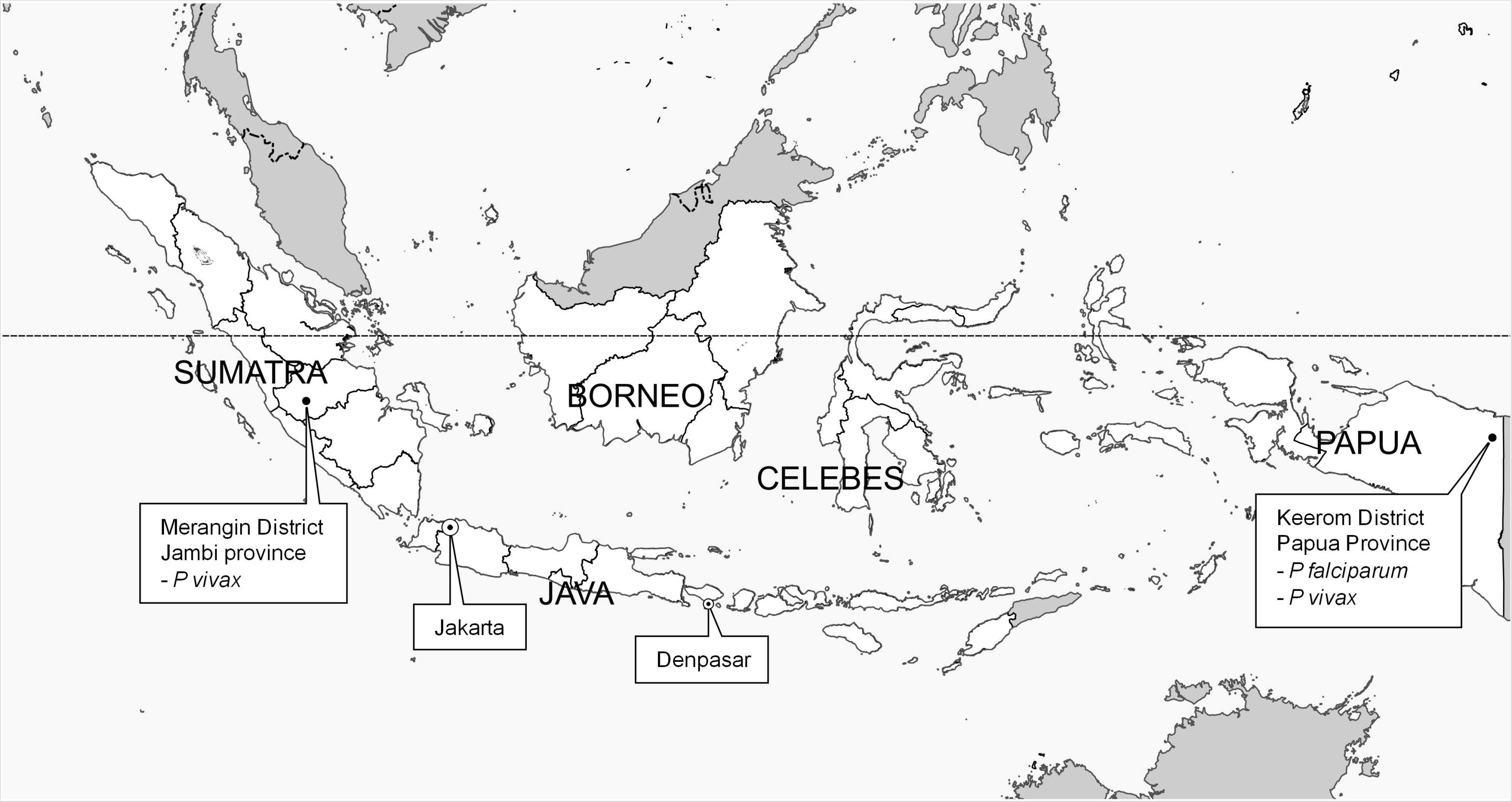
Study Sites in Papua and Jambi Provinces. Map from Natural Earth. https://www.naturalearthdata.com/

### *In vivo* drug efficacy study

Participants were recruited from among malaria-infected persons found during active malariometric surveys (active case detection (ACD)) or from persons attending outpatient clinics at Primary Health Centres (passive case detection (PCD)) in Keerom District and Merangin District. Persons were eligible to enrol in the study if they were aged between 1 - 65 years, weighed more than 5 kgs, had fever or history of fever in the preceding 24 hours, with slide-confirmed malaria with parasitemia of more that 500/ul asexual parasites for *P. falciparum* and more than 250/ul asexual parasites for *P. vivax*. Persons were excluded if they met any of the following exclusion criteria: 1) were pregnant, 2) had a history of allergy to the study drugs or study drug’s derivative, 3) had previously completed treatment with an antimalarial drug in the preceding two weeks, or 4) had a medical history of untreated hypertension or chronic heart, kidney, or liver disease [6].

### Laboratory procedures

Before enrolment, a finger prick was performed to obtain blood to prepare thick and thin blood smears and blots on filter paper (Whatman International Ltd., Maidstone, United Kingdom), for parasite genotyping, and for hemoglobin measurements (HemoCueTM Hb201+; HemoCue, Angelholm, Sweden). Smears and filter paper blood samples were also collected from finger pricks on days 1, 2, 3, 7, 14, 21, 28, 35 and 42 [6, 7]. Smears were read by expert microscopists and confirmed by polymerase chain reaction (PCR). Any discordant results between microscopy and PCR were resolved by independent PCR confirmation. All study participants had the G6PD level checked using Care start Rapid G6PD deficiency whole blood test.

### Antimalarial therapy

All study participants were given a supervised treatment of dihydroartemisinin (DHA) and piperaquine (PPQ), containing 40 mg DHA and 320 mg PPQ per tablet and was administered once a day for 3 days, as a weight per dose regimen of 2.25 and 18 mg/kg of DHA-PPQ [8] and followed-up weekly for 42 days. A study nurse distributed the drugs, observed and recorded all treatments, and repeated the treatment if vomiting occurred within 30 minutes following the drug administration. Parasitological responses were classified according to criteria of the World Health Organization [9]. Adverse events observed during the study were recorded by the study nurse and/or physician. Primaquine therapy was not provided until discontinuation from the study i.e., day of recurrence or day 42.

### Parasitological analysis

Thick and thin blood smears were stained with Giemsa and subsequently examined in light microscopy. Parasitemia was determined with parasite counted per 200 white blood cells in follow-up smears, counting will be done against at least 500 white blood cells. A blood slide will be considered negative when examination of 1000 white blood cells or 100 fields containing at least 10 white blood cells per field reveals no asexual parasites. All slides were read by a certified microscopist and cross-checked by a second experienced microscopist. In cases where readings were discordant, the slides were reread by third microscopist and a consensus reached.

### Preparation of genomic DNA

Parasite and human host DNA (on day of enrolment and day of recrudescence) was extracted from blood samples using Chelex-100 ion exchanger (Bio-Rad Laboratories, Hercules, CA) according to a previously published procedure [10]. Extracted DNA was either used immediately for Polymerase Chain Reaction (PCR) assays or stored at –20°C for later analysis.

### PCR Correction

Plasmodium was identified using microscopic test, which was followed by an evaluation using PCR. The PCR involved five primer sets. In the first amplification reaction (Nested 1), a pair of primers with genus specific of rPLU1 and rPLU5 was used and a total of 25 µl volume was used for all reactions. Furthermore, primers for the second amplification reaction (Nested 2) were used following the procedure described previously [11]. Data obtained from microscopy reading and PCR correction were calculated using Kaplan-Meier analysis.

### Genotyping and PCR amplification of *P. falciparum* and *P. vivax* genes

Genotyping using the genes for merozoite surface protein 1(MSP1), MSP2, and glutamate-rich protein (GLURP) was performed in certain participants to distinguish between pre-treatment and recrudescent parasites [12]. Amplifications of Pf K13 and Pv K12 genes for artemisinin resistance were performed according to previously published. The DNA was amplified by nested PCR and sequencing method to detect the mutations G449A, N458Y, T474I; M476I; A481V; Y493H; T508N; P527T; G533S; N537I; R539T; I543T; P553L; R561H; V568G; P574L; C580Y of *P. falciparum* K13 [13 – 17]. Identification of 8 nonsynonymous K12 mutations at codons M448, T517, F519, I568, S578, D605, D691, L708 for K12 of *P. vivax* K12 [18].

### Quantitative PCR to assess P. falciparum plasmepsin 2 gene copy number

Copy number of *P.falciparum* plasmepsin 2 gene determination consisted of several stages. Initially, DNA was extracted from the blood spots on filter paper according to the Wooden method [10] and purified using Qiagen Kit. The DNA extract were then used as templates in the amplification process of the copy number gene target *PfPM2* and Pftub genes using quantification of the real time polymerase chain reaction (RT-qPCR) and assay parameters according to the Witkowski method. The primers used for PfPM2 gene were 5’-TGGTGATGCAGAAGTTGGAG-3’ and 5’-TGGGACCCATAAATTAGCAGA-3’, while for Pftubulin these were 5’-TGATGTGCGCAAGTGATCC-3’ and 5’-TCCTTTGTGGACATTCTTCCTC-3’ [19]. Each control and samples were quantified in triplicates for PfPM2 and Pftub. The 3D7 strain were quantified in 6 replicates for PfPM2 and Pftub. Interpretation of results and run validation followed the Witkowski method. The 3D7 strain line was included in each run as standard control for one copy of PfPM2 gene in 6 replicates. PfPM2 copy number was calculated by the 2-ΔΔCt method [19] and the value was rounded up.

## RESULTS

### Keerom District, Papua

Of the 6339 subjects screened through passive and active case detections (Fig 2), 1984 (31.3%) were found positive for malaria. Falciparum malaria dominated the malaria cases (56%, 1112/1984) and followed by vivax malaria at 37.8% (749/1984). An additional 3.4% (68/1984) were positive for *Plasmodium malariae*; 1 (0.05%) had *Plasmodium ovale*; 42 (2.1%) had mixed infections of *P. falciparum* and *P. vivax, P. falciparum* and *P. ovale* 1 (0.05%), 1 (0.05%) *P. vivax* and *P. malariae* (Fig 2 – 3).

**Figure 2.**
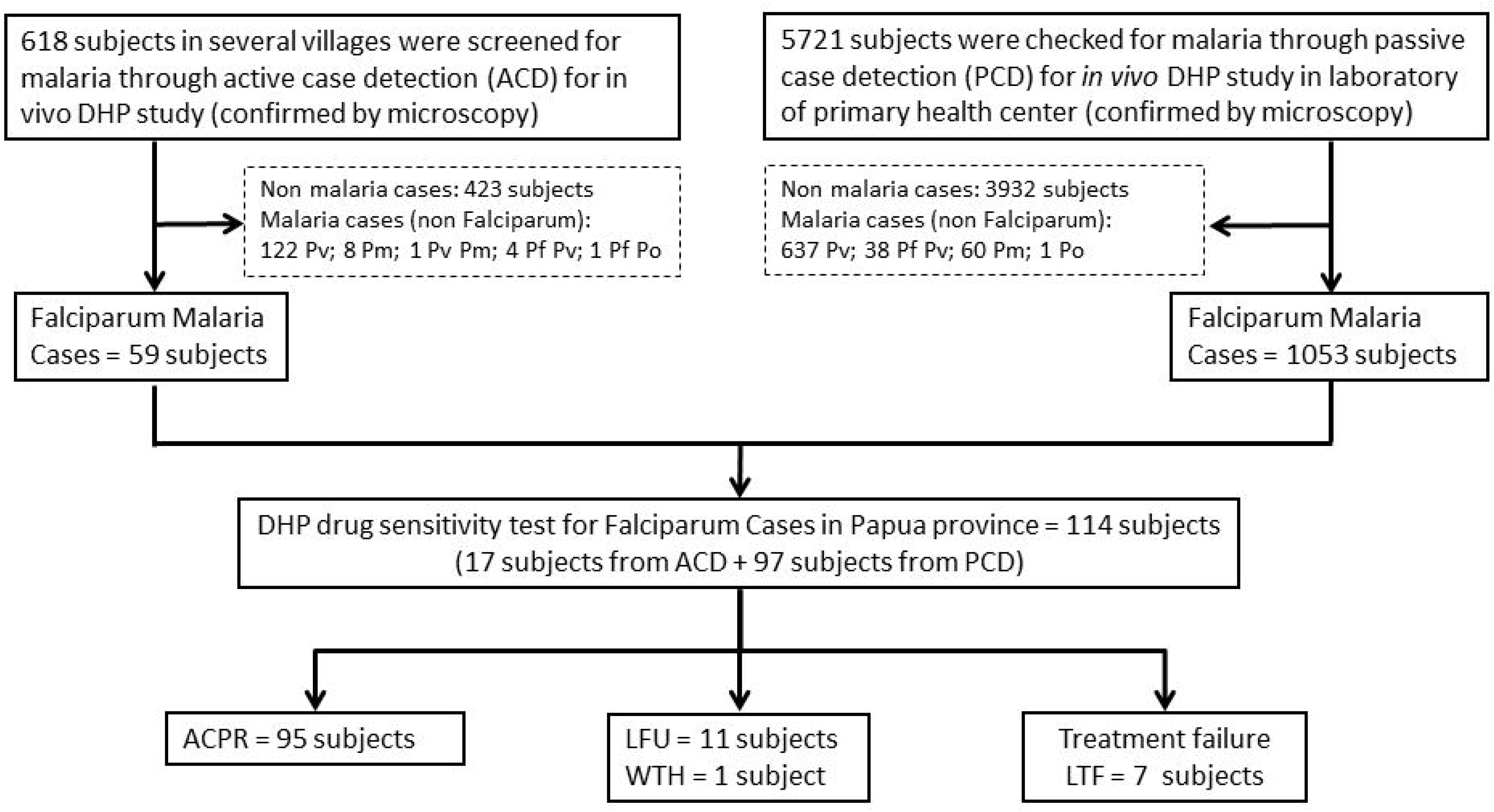
Flow chart sample collection for falciparum cases in Keerom, Papua

A total of 114 (5.7%) of the 1984 *P. falciparum* cases and 83 (4.2%) *P. vivax* cases met inclusion criteria (Table 1; Fig 2–3). The remaining subjects were excluded due to age, inadequate asexual parasitemia, unplanned travelling, refusal to provide consent, and local tribes war (unsecure situation for follow up activity).

**Figure 3.**
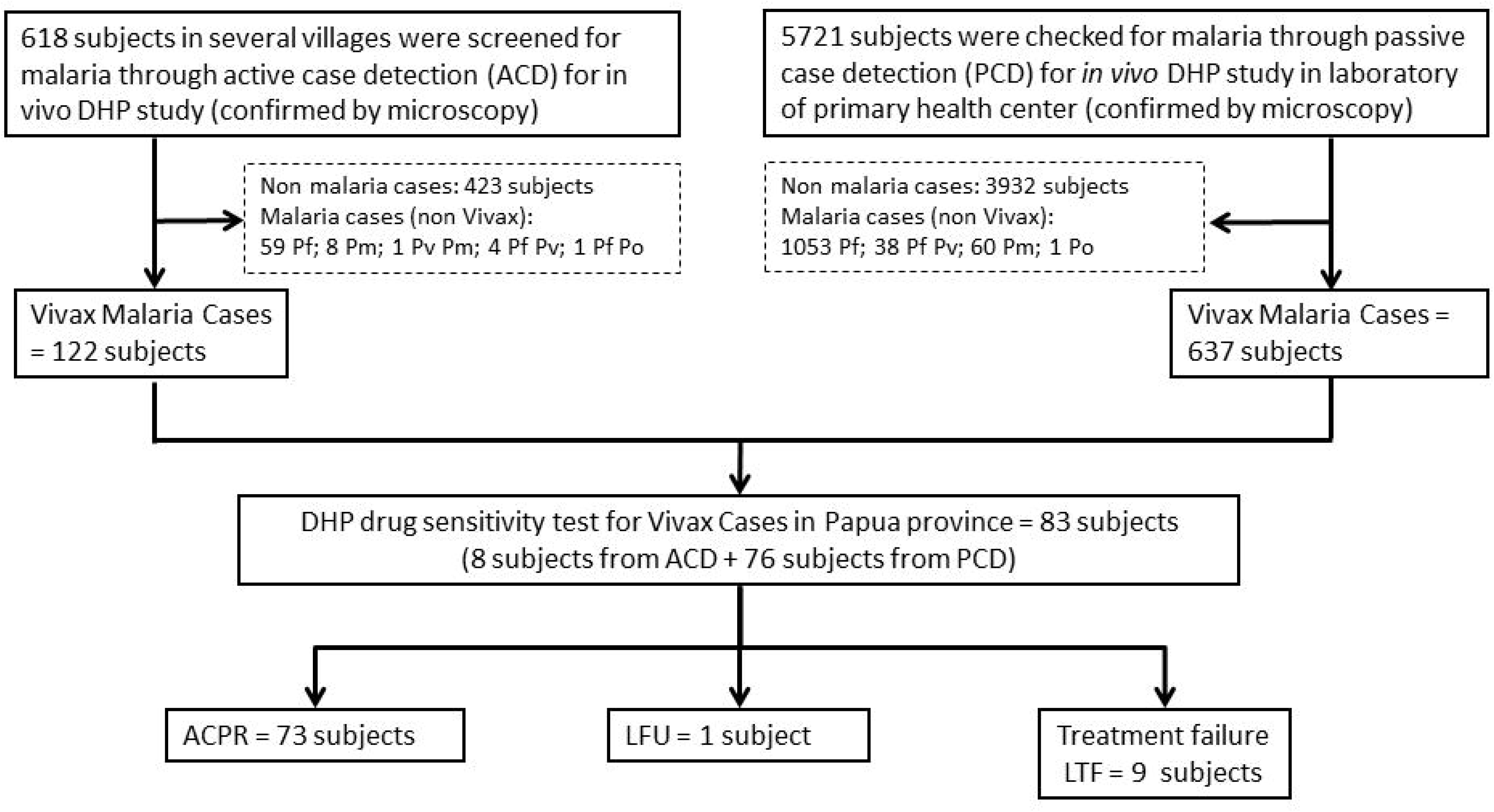
Flow chart sample collection for vivax cases in Keerom, Papua

**Table 1.**
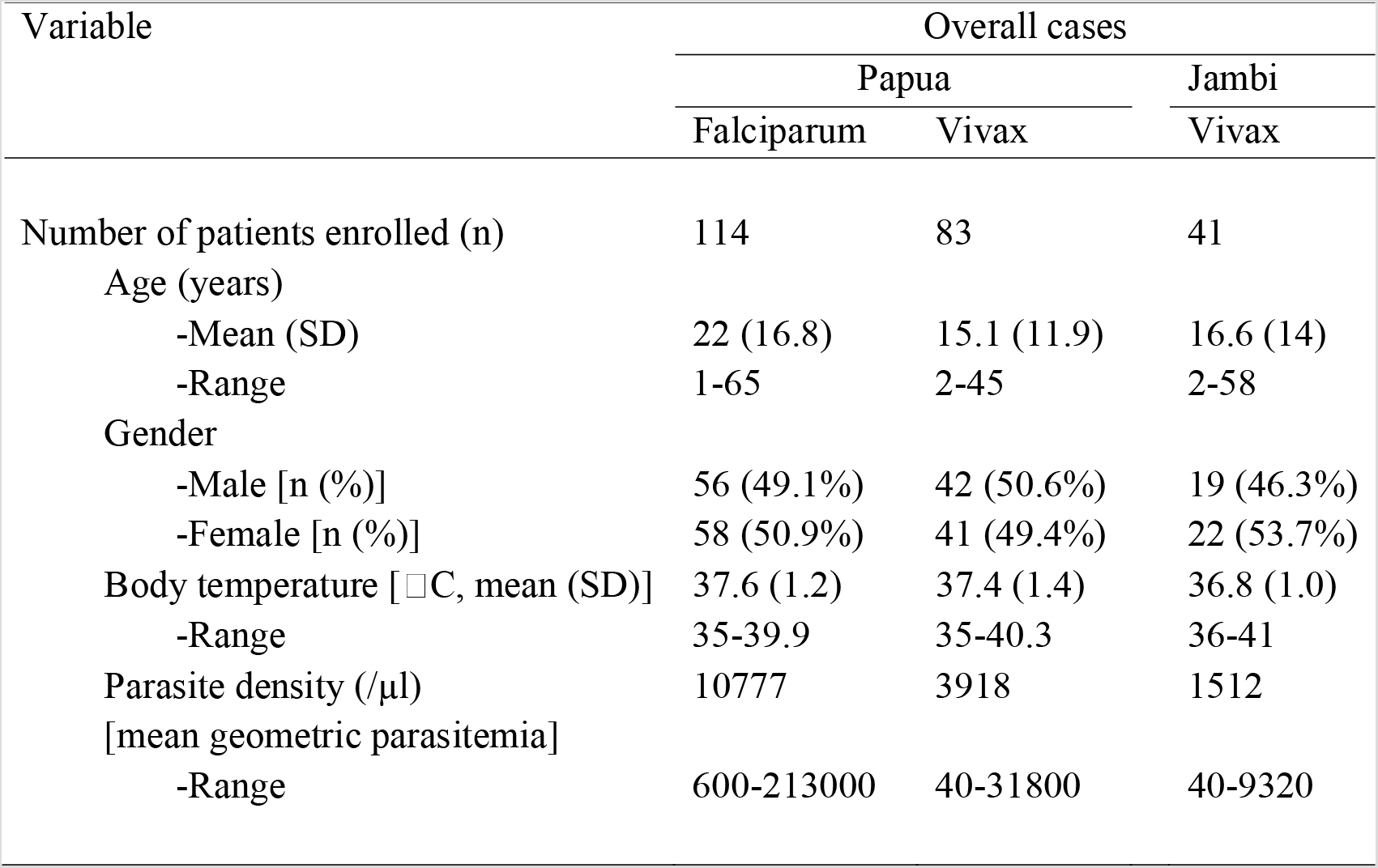
Baseline characteristics of study participants

| Variable | Overall cases |  |  |
| --- | --- | --- | --- |
|  | Papua |  | Jambi |
|  | Falciparum | Vivax | Vivax |
| Number of patients enrolled (n) | 114 | 83 | 41 |
| Age (years) |  |  |  |
| -Mean (SD) | 22 (16.8) | 15.1 (11.9) | 16.6 (14) |
| -Range | 1-65 | 2-45 | 2-58 |
| Gender |  |  |  |
| -Male [n (%)] | 56 (49.1%) | 42 (50.6%) | 19 (46.3%) |
| -Female [n (%)] | 58 (50.9%) | 41 (49.4%) | 22 (53.7%) |
| Body temperature [°C, mean (SD)] | 37.6 (1.2) | 37.4 (1.4) | 36.8 (1.0) |
| -Range | 35-39.9 | 35-40.3 | 36-41 |
| Parasite density (/µl) | 10777 | 3918 | 1512 |
| [mean geometric parasitemia] |  |  |  |
| -Range | 600-213000 | 40-31800 | 40-9320 |

Table 1 lists the demographic characteristics of the study subjects at enrolment. Of the 114 enrolled subjects for falciparum cases, 56 were males and 58 were females with age ranging from 1 to 65 years (mean 22 years). At enrolment, the density of asexual forms ranged from 600 to 213000 per µl blood, whereas sexual stages (gametocytes) were found in 5 subjects (4.4%) (Table 7). During the follow up, gametocytes were found in few cases (Table 4–5).

### Clinical and parasitological efficacy of DHA-PPQ for falciparum cases in Keerom, Papua

Classification of the treatment outcomes (microscopy and PCR corrected) for falciparum cases is presented in Table 2. At day 42, ACPR was noted in 93.1% by microscopy and PCR correction. No patients showed ETF, while LCF was reported in 1 study participant. LPF was observed in 6 study participants with PCR correction. Withdrawn or drop out was observed in 1 case (0.9%). Lost to followed up was observed in 11 study participants. Of the 114 falciparum cases enrolled, 112 cases were successfully cleared at day 2. The remaining 2 cases were cleared at day 3. No delayed parasite clearance was observed. Recurrent parasites were detected in 6 cases, at days 21, 35 and day 42 (Table 6).

**Table 2.** Falciparum phenotyping result from Keerom, Papua during the 42 days of follow up

| Study Site | Total Subject | Diagnosis by | Outcome (Number of patients) |  |  |  |  |  |
| --- | --- | --- | --- | --- | --- | --- | --- | --- |
|  |  |  | ETF | LCF | LPF | ACPR | LTU | WTH |
| Papua | 114 | Microscopy | 0 | 1<br>(1%) | 6<br>(5.9%) | 95<br>(93.1%)<br>(95%CI:86.4-97.2) * | 11 | 1 |
|  | 114 | PCR corrected | 0 | 1<br>(1%) | 1<br>(1%) | 95<br>(97.9%)<br>(95%CI:92.7-99.7) | 11 | 6 |

ETF = Early treatment failure; LCP = Late clinical failure; LPF = Late parasitological failure; LTU = Lost to follow up; WTH = Withdrawn ACPR = Adequate clinical and parasitological response; (\*) = Kaplan-Meier Analysis

### Differentiation of recrudescent with reinfection

Genotypic analyses of the parasites at day 0 and day of recurrence were conducted using the 3-markers recommended by WHO; *msp1, msp2*, and *glurp* genes as shown in Table 6. Of the 6 LPF cases, 4 cases were categorized as re-infection as the genotypes of the parasites found at day of recurrence were different with that of day 0 (pre-treatment). The remaining 2 subjects showed the same genotypes for the 3 markers and therefore could be determined as either recurrence or re-infection. Nonetheless, either case indicate resistance as they have survived the challenge of sub-curative dose of piperaquine

### Determination of the existence of SNPs in the PfK13

PCR amplification and DNA sequencing of the Pf K13 gene to observe the 20 SNPs associated with Artemisinin resistance; G449A, N458Y, T474I; M476I; A481V; Y493H; T508N; P527T; G533S; N537I; R539T; I543T; P553L; R561H; V568G; P574L; C580Y revealed that all *P. falciparum* isolates carried the wildtype allele.

### *Plasmodium falciparum* plasmepsin 2 gene copy number

Late parasitological failure (LPF) was observed in 6 study participants with microscopy reading (Table 2). The delta Ct from 6 LPF were compare with Ct from control, *P. falciparum* strain 3D7. The estimation of the copy number from 6 LPF was calculated (Table 7). Three LPF (PAF 01, 08, and 19) had the same copy number of plasmepsin 2 in day 0 and day recurrence. Two LPF (PAF 37 and 112) have increased 2 copy number of plasmepsin 2 in day 0 and day recurrence, while 1 LPF (PAF 133) had copy number 3. Of the 6 recurrent *P. falciparum* found, 2 indicated recrudescence and 4 cases were re-infections (Table 9). Analysis of the PfPM2 gene at day 0 and day of recurrence in recrudescent cases (PAF 01 and 19) revealed the same single copy number, whereas 3 of the 4 re-infection cases carried 2-3 copy numbers (Table 7).

### Clinical and parasitological efficacy of DHP for vivax cases in Keerom, Papua

Table 3 lists the demographic characteristics of the study subject at enrolment. Of the 83 enrolled subjects for vivax cases, 42 were males and 41 were females with age ranging from 2 to 45 years (mean 15 years) (Table 1). At enrolment, the density of asexual forms ranged from 40 to 31800 µl, whereas the sexual stages (gametocytes) were found in 15 subjects (18.1%) (Table 8). At follow up, gametocytes were only found in 3 cases; 1 in day 2, 1 in day 3 and 1 in day 42 (Table 5). Classification of the treatment outcomes (microscopy and PCR corrected) for vivax cases is presented in Table 6. At day 42 an ACPR was noted in 89% by microscopy and 100% after PCR correction. No patients showed ETF for vivax cases, while LCF was reported in 1.2%. LPF was observed in 8 study participants without PCR correction. Lost to followed up was observed in 1 study participant (Table 3). Of the 83 vivax cases enrolled, no delayed parasite clearance was observed.

**Table 3.** Vivax phenotyping result from Keerom, Papua and Merangin, Jambi Provinces during the 42 days of follow up

| Study Site | Total Subject | Diagnosis by | Outcome (Number of patients) |  |  |  |  |  |
| --- | --- | --- | --- | --- | --- | --- | --- | --- |
|  |  |  | ETF | LCF | LPF | ACPR | LTU | WTH |
| Papua | 83 | Microscopy | 0 | 1<br>(1.2%) | 8<br>(9.8%) | 73<br>(89%)<br>(95%CI:80.2-94.9) * | 1 | 0 |
|  | 79 | PCR corrected | 0 | 0 | 0 | 73<br>(100%)<br>(95%CI:95.1-100) | 1 | 5 |
| Jambi | 41 | Microscopy | 0 | 0 | 1<br>(2.6%) | 37<br>(97.4%)<br>(95%CI:86.2-99.9) | 2 | 1 |
|  | 40 | PCR corrected | 0 | 0 | 0 | 37<br>(100%)<br>(95%CI:90.5-100) | 2 | 1 |

ETF = Early treatment failure; LCP = Late clinical failure;
LPF = Late parasitological failure; LTU = Lost to follow up; WTH = Withdrawn
ACPR = Adequate clinical and parasitological response; (\*) = Kaplan-Meier Analysis

**Table 4.**
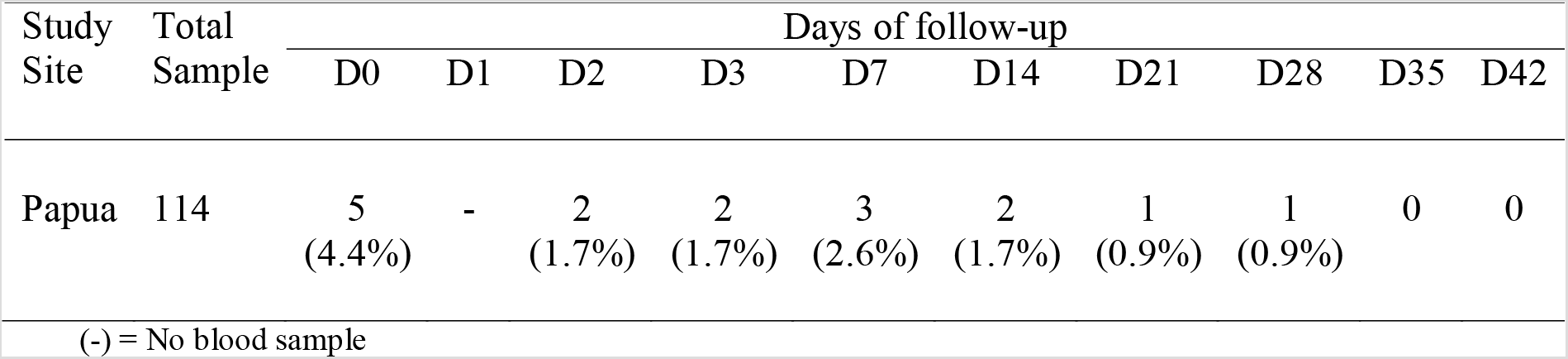
Gametocyte of *P. falciparum* appearance during the 42 days of follow up

| Study Site | Total Sample | Days of follow-up |  |  |  |  |  |  |  |  |  |
| --- | --- | --- | --- | --- | --- | --- | --- | --- | --- | --- | --- |
|  |  | D0 | D1 | D2 | D3 | D7 | D14 | D21 | D28 | D35 | D42 |
| Papua | 114 | 5<br>(4.4%) | - | 2<br>(1.7%) | 2<br>(1.7%) | 3<br>(2.6%) | 2<br>(1.7%) | 1<br>(0.9%) | 1<br>(0.9%) | 0 | 0 |

(-) = No blood sample

### Merangin District, Jambi Province

Of the 751 subjects screened through passive and active case detection, 50 (66.7%) were found positive for malaria. Only vivax malaria cases were found (Fig 4). A total of 41 (82%) *P. vivax* cases met inclusion criteria. The remaining subjects were excluded due to age, inadequate asexual parasitemia, and refusal to provide consent. The demographic characteristics of the study subject at enrolment is shown in Table 1. Of the 41 enrolled subjects for vivax cases, 19 were males and 22 were females with age ranging from 2 to 58 years (mean 16 years). At enrolment, the density of asexual forms ranged from 40 to 9320 µl, whereas sexual stages (gametocytes) were found in 18 subjects (43.9%) (Table 5).

**Figure 4.**
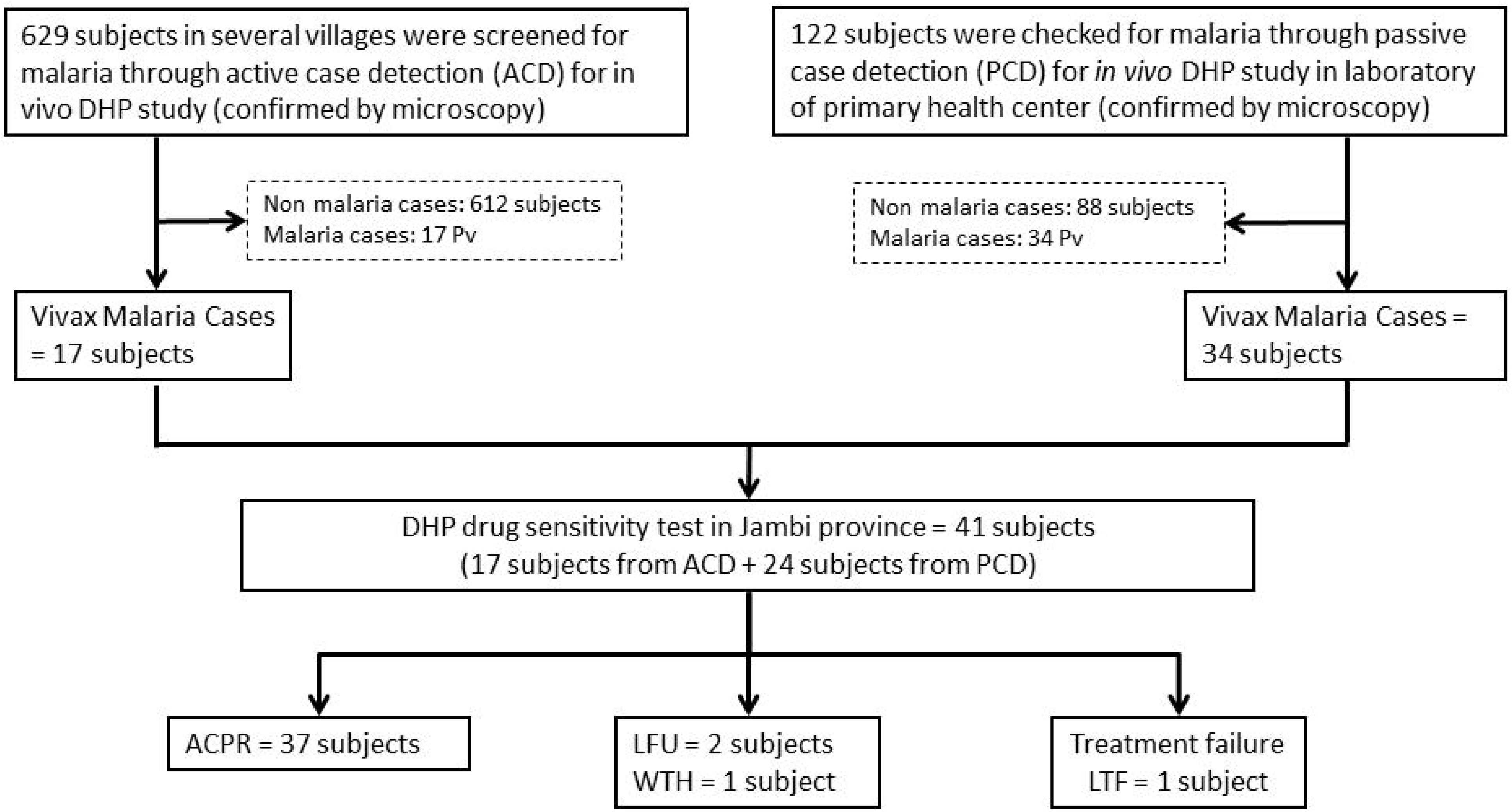
Flow chart sample collection for vivax cases in Merangin, Jambi

**Table 5.** Gametocyte of *P. vivax* appearance during the 42 days of follow up

| Study Site | Total Sample | Days of follow-up |  |  |  |  |  |  |  |  |  |
| --- | --- | --- | --- | --- | --- | --- | --- | --- | --- | --- | --- |
|  |  | D0 | D1 | D2 | D3 | D7 | D14 | D21 | D28 | D35 | D42 |
| Papua | 83 | 15<br>(18.1%) | - | 1<br>(1.2%) | 1<br>(1.2%) | 0 | 0 | 0 | 0 | 0 | 1<br>(1.2%) |
| Jambi | 41 | 18<br>(43.9%) | - | 0 | 0 | 0 | 0 | 0 | 0 | 1<br>(2.4%) | 0 |

(-) = no blood sample

### Clinical and parasitological efficacy of DHP for vivax cases in Merangin, Jambi

Classification of the treatment outcomes (microscopy and PCR corrected) for vivax cases is presented in Table 3. At day 42, ACPR was noted in 97.4% of the cases by microscopy and 100% after PCR correction. No patients showed ETF and LCF for vivax cases, while LPF was reported in 2.6%. LPF was observed in 1 study participant without PCR correction. One study participant was withdrawn due to unscheduled traveling. Lost to followed up was observed in 2 study participants (Table 3). No gametocyte was found after the treatment completed and during the follow up period up to day 42 except for 1 case where gametocyte appeared at day 35.

### Determination of the existence of SNPs in the PvK12

PCR amplification and DNA sequencing of the Pv K12 gene to observe the 8 SNPs associated with Artemisinin resistance; M448, T517, F519, I568, S578, D605, D691, L708 revealed that all *P. vivax* isolates carried the wildtype allele for samples from Keerom District and Merangin District.

### Evaluation of the adverse event

No adverse event was reported during the follow up of this study in Keerom (Papua) and Merangin (Jambi).

### Gametocyte carriage during the treatment

Gametocytemia was present at enrolment in 5 patients in Keerom District (Papua) infected with *P. falciparum* and 15 patients with *P. vivax* malaria while 18 patients had gametocyte carriage at day of enrollment in Merangin District (Jambi) (Table 5). The proportion of patients with patent gametocytemia in those with *P. falciparum* infection was 4.4% at D0, 1.7% at D2, 1,7% at D3, and 2.6% at D7. In Keerom patients with *P. vivax* malaria, the proportion with gametocyte fell from 18.1% at D0 to1.2% at D2, D3 and D42, while in Merangin District (Jambi) the proportion of patients with patent gametocytemia in those with

*P. vivax* infection was 43.9% at D0 and 2.4% at D35 (Table 5).

### Discussion

Development and spread of the parasite resistance to the currently available artemisinin-based combination therapy (ACT) poses a substantial threat to the currently endorsed malaria elimination program as it may increase not only malaria morbidity but also re-introduction of malaria in areas where elimination have been achieved. Results of this study clearly indicate that DHA-PPQ is still highly effective in both study sites. Nevertheless, in Keerom District Papua, evidence for the existence of parasite isolates that are resistant to PPQ alerts to the proper deployment of the drug in the area. Piperaquine resistance is associated with the increased copy number of the plasmepsin gene [19, 20] and as a result the treatment failed to completely eliminate the parasite from the blood or prevent re-infection during the follow up period. This study found the presence of 2 recrudescent cases at days 21 and 35 and reinfection at days 35-42 (Table 6). Recrudescence and re-infection following treatment with DHP were associated with higher prevalence of Kelch13 mutations, higher piperaquine 50% inhibitory concentration (IC50) values, and lower mefloquine IC50 values [19 – 23]. The high gametocyte carriage at enrolment in vivax cases (∼43.9%) in this study (Table 5) is likely associated with the poor accessibility to treatment and also compliance to the treatment regimen. Almost 50% of the subjects had received previous DHA-PPQ but never completed the 14 day primaquine treatment as recommended. This situation requires attention as it may expedite parasite resistance to the DHA-PPQ as well as support for local transmission. In Papua the efficacy of DHA-PPQ against *P. falciparum* and *P. vivax* were 97.9% and 100% respectively, with 6 recurrent infections for falciparum malaria. However, the proportion of parasitemic patients fell rapidly and none of the patients were parasitemic at day 3. This findings indicate that artemisinin is still highly effective but caution has to be given to the partner drug, piperaquine. The results of the molecular analysis also support the finding as none of the parasites had the polymorphisms in the K13 and K12 gene that have previously been associated with artemisinin resistance. However, of the 6 recurrent parasites, 3 carried the amplification of the *plasmepsin* 2-3 gene cluster. The evidence for DHA-PPQ late treatment failure in this study alerts to the proper treatment of malaria in the area and also anticipate having second line ACT to replace the piperaquine partner drug. Currently, the Indonesia national policy to use quinine as second line drug is regarded to be impractical as it introduces longer treatment period and also more often side effect. In this regard, consideration of using another ACT such as Arthemeter+lumefantrine or Artesunate+mefloquine at least for falciparum cases might be rational. The results from this study are reassuring and suggest that in the absence of artemisinin resistance, the ACT regimen may delay de novo emergence of resistance to the partner drug. DHA-PPQ has been used in Indonesia as the first line antimalarial drug since 2008 [24] and it took almost 10 years to first detect the early sign of resistance to the partner drug, piperaquine. In Cambodia, amplification of the *plasmepsin* 2-3 gene cluster has been identified as an important molecular determinant of piperaquine resistance in *P. falciparum* [21 – 23] and resistance to piperaquine in fact increase the sensitivity of the mefloquine [19]. This phenomenon may complement for the replacement of piperaquine as partner drug of artemisinin in the area if the resistance is spread. In Indonesia, DHA-PPQ procurement is highly regulated by the Indonesian Ministry of Health, and the drug is only available at government health facilities and selected private-sector facilities, which are able to confirm that the prescription should be based on malaria positivity by microscopy or rapid diagnostic test. With this tight regulation it is anticipated that DHA-PPQ will continue to play a role in the treatment of uncomplicated malaria in Indonesia until malaria is successfully eliminated in the country. On another aspect, implementation of evidence-based vector control may also contribute to mitigate transmission and delay the emergence of antimalarial drug resistance.

**Table 6.**
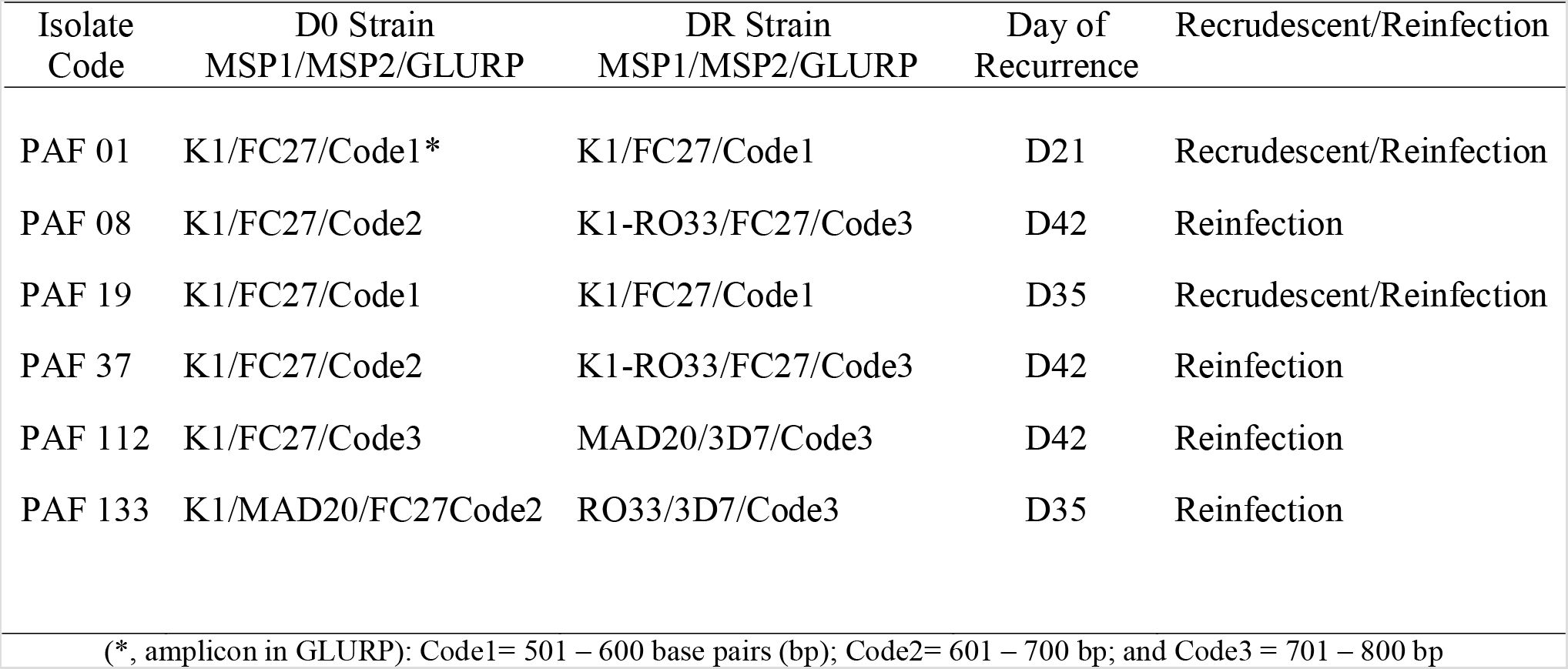
Genotyping results of the parasites at day 0 and day recurrence in *P. falciparum* cases solate Code

**Table 7.**
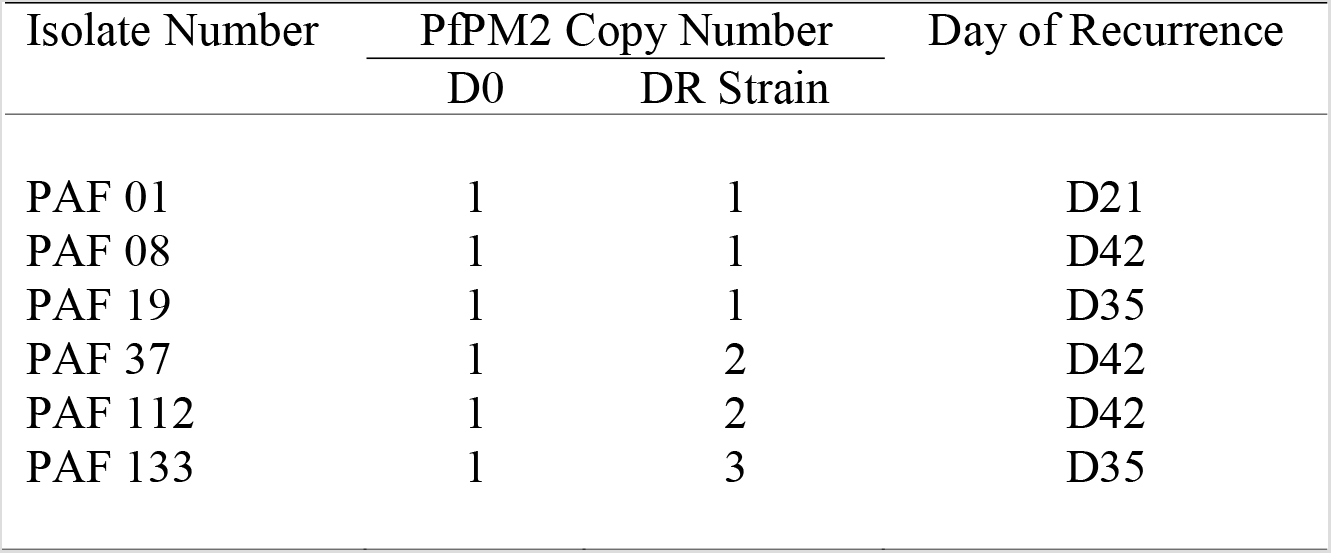
PfPM2 Gene Copy Number of the parasites at day 0 and day recurrences in *P. falciparum* cases

### Conclusions

The therapeutic efficacy study conducted in two sentinel sites in Papua and Jambi, Indonesia during 2017-2018 revealed that DHA-PPQ is still highly effective in both sites. The appearance of recurrent falciparum infection in small number of cases in Papua alert to the possible emergence of piperaquine resistance in the area and deserve further investigation to contain its spread and anticipate for the rational option of a second line ACT. Further study is required in different regencies in Papua, particularly those in border area with Papua New Guinea to determine the spread of resistance to DHA-PPQ.

## Data Availability

All relevant data are within the manuscript

## Acknowledgments

The authors are grateful to participants in this study. The authors are deeply grateful to Chairman of the Eijkman Institute for Molecular Biology (EIMB), Professor Amin Soebandrio MD., Ph.D., Clin. Microbiol and Coordinator Director Office Global Malaria Programme WHO, Dr. Pascal Ringwald for their support and encouragement in this activity. The authors wish to thank Ahmad N Azhari from WHO, staffs from Papua and Jambi Provincial Health Departments, Keerom and Merangin District Health Departments and Primary Health Centers at study sites for the support, encouragement and helping for samples collection. We thank Nadha Rizky Pratama, Sylvia Sance Marantina, Jenifer Kiem Aviani, and Annisa Rizkia for their assistance in the EIMB laboratory.

## Data availability statement

All relevant data are within the manuscript

## Author Contribution

Conceptualization: Maria Dorina G Bustos, Din Syafruddin

Data curation: Puji BS Asih, Ismail E Rozi, Farahana K Dewayanti, Suradi Wangsamuda, Syarifah Zulfah, Marthen Robaha, Jonny Hutahaean, Nancy D Anggraeni, Marti Kusumaningsih, Pranti S Mulyani, Elvieda Sariwati, Herdiana H Basri,

Formal analysis: Puji BS Asih, Ismail E Rozi, Farahana K Dewayanti, Suradi Wangsamuda, Maria Dorina G Bustos, Din Syafruddin

Funding acquisitions: Maria Dorina G Bustos, Din Syafruddin

Investigation: Puji BS Asih, Ismail E Rozi, Farahana K Dewayanti, Suradi Wangsamuda, Maria Dorina G Bustos, Din Syafruddin

Methodology: Puji BS Asih, Ismail E Rozi, Farahana K Dewayanti, Maria Dorina G Bustos, Din Syafruddin

Project administration: Puji BS Asih, Syarifah Zulfah, Marthen Robaha, Din Syafruddin Supervision: Marti Kusumaningsih, Pranti S Mulyani, Syarifah Zulfah, Marthen Robaha, Maria Dorina G Bustos, Din Syafruddin

Writing – original draft: Puji BS Asih, Din Syafruddin

Writing – review & editing: Puji BS Asih, Ismail E Rozi, Farahana K Dewayanti, Suradi Wangsamuda, Syarifah Zulfah, Marthen Robaha, Jonny Hutahaean, Nancy D Anggraeni, Marti Kusumaningsih, Pranti S Mulyani, Elvieda Sariwati, Herdiana H Basri, Maria Dorina G Bustos, Din Syafruddin

## Author Summary

This study aims to determine the efficacy and safety of dihydroartemisinin-piperaquine (DHA-PPQ) to treat malaria patients in Indonesia. The study was conducted in 2 sites, Keerom, Papua and Merangin, Jambi Provinces. In Keerom District, a total of 114 *P. falciparum-*infected and 83 *P. vivax*-infected were recruited, treated under supervision with DHA-PPQ once daily for 3 days. Kaplan-Meier analysis of microscopy readings and PCR-corrected falciparum cases revealed a 93.1% and 97.9% efficacy, respectively and were classified as Adequate Clinical Parasitological Responses (ACPRs). For vivax malaria, the DHA-PPQ efficacy were 89% and 100%. In Merangin District, 751 subjects were screened and 41 subjects were recruited. Microscopy reading and PCR-corrected analysis revealed a 97.4% and 100% efficacy. No severe adverse events were found in both sites. No delay in parasite clearance was found and no mutations observed in the PfK13 and PvK12 genes. Of the 6 recurrent *P. falciparum* found, 2 indicated recrudescent and 4 cases were re-infection. Analysis of the PfPM2 gene at day 0 and day of recurrence in recrudescent cases revealed the same single copy number, whereas 3 of the 4 re-infection cases carried 2-3 copy numbers. Treatment of falciparum and vivax malaria cases with DHA-PPQ showed a high efficacy and safety.

